# Characterization of maternal mortality and severe maternal morbidity before and during covid-19: an analysis for the Dominican Republic

**DOI:** 10.1101/2024.03.11.24304148

**Authors:** Kary Desiree Santos-Mercedes, Raquel Zanatta Coutinho

## Abstract

The Covid-19 pandemic disturbed the delivery of health services, which included obstetric care, in many parts of the word. In countries where maternal mortality was already elevated before the pandemic, this disruption brought about catastrophic events. Using data from the Sistema Nacional de Vigilancia Epidemiológica (SINAVE) of the maternal morbidity and mortality of the Dominican Republic, this paper estimated the severe maternal morbidity rate and the maternal mortality rate by causes of death (direct, indirect, and specific causes). Mixed effects models were used to identify individual and contextual factors that contribute to a higher risk of dying. Results indicate that the severe maternal morbidity rate decrease from 12.1 to 10.4 per 1,000 live births in 2020 compared to 2019; however, the maternal mortality rate went from 91.4 to 124.2 per 100,000 live births in the same period. In 2021, a significant increase in morbidity and mortality was observed, registering a rate of 16.5 cases of severe maternal morbidity per 1,000 live births and a rate of 153.7 maternal deaths per 100,000 live births. Additionally, maternal mortality in the Dominican Republic is associated with the sociodemographic and economic conditions of women, revealing inequalities related to national origin, area of residence and access to health services which were worsening during the pandemic.

## INTRODUCTION

Maternal mortality is an indicator closely related to the economic and social development of countries, revealing inequalities in women’s living conditions. In developing countries, the risk of maternal death over a woman’s lifetime is nearly 40 times higher than in developed countries. Approximately 830 women worldwide die each day from pregnancy- or childbirth-related complications (1). Almost all of these deaths occur in low-income countries (1,2). In Latin America and the Caribbean, approximately 15,000 women die each year from pregnancy-related causes (3,4). Research shows that two-thirds of maternal deaths are preventable with timely medical care and ideal conditions (5).

Maternal death is defined as the death of a woman during pregnancy or within 42 days after childbirth or termination of pregnancy, regardless of the duration or location, due to any pregnancy-related cause aggravated by pregnancy or its management, but not by accidental or incidental causes (6). WHO uses the term Severe Maternal Morbidity (SMM) to describe obstetric events occurring during pregnancy, childbirth, or the 42 days postpartum, whether acute, chronic, or both, in which the mother survived but was at risk of dying (1). It is estimated that these women share the same pathological and circumstantial conditions as those who actually died from the same complication, providing better information about the entire process(4).

Despite the high number, maternal survival has significantly improved since the adoption of the Sustainable Development Goals. Between 1990 and 2013, global maternal mortality decreased by 45%. And more than 71% of births worldwide were attended by skilled health personnel in 2014, compared to around 59% in 1990. However, maternal mortality remains high in some parts of the world, reflecting inequalities in access to health care services and highlighting the gaps between rich and poor, reflecting social inequalities, low coverage, and poor quality of health services (7).

The maternal mortality ratio in developing countries in 2015 was 239 maternal deaths per 100,000 live births, compared to developed countries which had only 12 maternal deaths per 100,000 live births. There are significant disparities between countries, but there are also disparities within countries, among women with high and low incomes, by educational level, and by location, whether urban or rural (1,4).

In 2013, the Dominican Republic showed a maternal mortality ratio of approximately 102 deaths per 100,000 live births, and by the year 2016, it had decreased to a ratio of 92.1 deaths per 100,000 live births. This indicator hides the heterogeneity of the country’s health situation, as there are health regions that had a higher maternal mortality ratio than the national average and others below it, such as the case of the Northwest Cibao Region (Indicator well above the average) and the Central Cibao Region (Indicator below the average) (8,9). These data exceed the average for the Latin American and Caribbean region (9). Despite governmental efforts in the matter, the Dominican Republic has not been able to experience a significant reduction in maternal mortality, which distances the country from achieving the 2030 Agenda and the Sustainable Development Goals (10,11). It is imperative to highlight that the Dominican Republic, being a country with a high middle income (8,476.75 USD per capita (12,13) that has a high coverage of prenatal care (98.9%) and institutional delivery (97.5%).

The COVID-19 pandemic caused an increase in maternal mortality in the Dominican Republic (13). Data shows that in 2020, a rate of 127 per 100,000 live births was recorded, well above the average for Latin America, which was 67 per 100,000 live births. Studies conducted during the first year of the COVID-19 pandemic show that pregnancy and the postpartum period could indeed pose additional risks for both women and babies (14). Still, high-income countries have not observed an increase in pregnancy-related mortality, leading to the belief that the increase in maternal mortality ratio due to COVID-19 could be explained by some situations related to the more vulnerable context and obstetric care directly affecting women and babies. According to (15), poor quality prenatal care, insufficient resources to manage emergency and critical care, racial disparities in access to maternity services, obstetric violence, and the pandemic pose additional barriers to accessing health care.

With the aim of understanding the situation of severe maternal morbidity and maternal mortality in the national context, characterizing maternal mortality due to Covid-19 in the Dominican Republic in individual, contextual, and healthcare system aspects, this work utilizes data from the National Epidemiological Surveillance System (SINAVE) and the online live births database for the period 2015-2021. First, severe maternal morbidity ratios (SMMR) were estimated for the period 2015-2021. Then, maternal mortality ratios (MMR) were estimated for the period 2015-2021 by cause of death (direct, indirect, and specific ICD causes), verifying if, from 2020 onwards, there was a change in the observed behavior for the period. Finally, sociodemographic inequalities to which pregnant women were exposed before and during the Covid-19 pandemic are examined from the perspective of social determinants of health, identifying individual and contextual factors that would contribute to a higher risk of dying from COVID-19.

### Literature review

The estimated fertility rate for the Dominican Republic was 2.2 (16). With a life expectancy at birth for 2021 of 74.02 years and infant mortality of 20.2 per 1,000 live births (17). The country had 21% of the population below the poverty line in 2019 (16,18). Between 2002 and 2014, there was a decrease in the percentage of pregnant individuals who attended four or more prenatal care visits during pregnancy, as it decreased from 93.5% to 92.9%. On the other hand, 99.7% of births were attended by skilled personnel in 2018 (19,20). Regarding COVID-19 in 2020, the Dominican Republic ranked 26th within the Americas Region in terms of the number of deaths from COVID-19 (16,18).

The Dominican state has undertaken various interventions in maternal and child health over the past decades with objectives focused on strengthening programmatic networks and collective health actions as established in international conferences of which the country was a signatory (1,21). Although the interventions have contributed to sustained reductions in maternal and child mortality, the program goals have not yet been achieved (2,10,22), such as the goal set in the MDG 5 to reduce by 75% the maternal mortality rate that existed in the early 1990s.

Santos and Patricio (8) argue that the national average tends to conceal the heterogeneity of the health situation within the country, as mortality assumes a differential behavior according to the geographical regions of each country. At the provincial level, the areas with the highest maternal mortality ratio for 2016 correspond, to a greater extent, to small provinces in terms of population, such as Independencia, Santiago Rodríguez, Bahoruco, Pedernales, and Elías Piña. Some of these provinces are located in the border area with Haiti, with high levels of poverty and limited coverage of some basic services. The results also show that the provinces of María Trinidad Sánchez and San Pedro de Macorís are in the group with the highest mortality ratio (8).

### Causes of maternal mortality

The International Statistical Classification of Diseases (ICD) typifies and defines deaths during pregnancy, childbirth, and the puerperium as direct or indirect maternal deaths (23). Direct maternal death is the consequence of complications due to pregnancy, childbirth, and the puerperium or their management. In 2016, in the Dominican Republic, they represented 74% of maternal deaths, with hypertensive disorders standing out as the leading cause of maternal death within this group (32%), followed by hemorrhages (11%), sepsis (10%), pregnancy terminated by abortion (8%), and complications of the puerperium (7%). Conversely, indirect maternal death is one generated as a consequence of pre-existing diseases or other associated conditions that appear during pregnancy and, although not related to it, lead to death because they are aggravated by the physiological effect of pregnancy. In 2016, they represented 26% of deaths. Examples of indirect deaths include malaria, hepatitis, or HIV infection (24). Additionally, late maternal death is classified as those occurring after 40 days of the puerperium but before one year after childbirth.

Regarding severe maternal morbidity, rates range from 3.8 to 12 per 1,000 births in developed countries. There have been few reports in Latin America, so the true extent of the problem is unknown, and studies have only been conducted in Brazil and Cuba (4,25–27). In Brazil, for example, they estimated that severe maternal morbidity is close to 21.13 cases per 1,000 admissions for the period 2010-2019 at the national level.

Various national and international studies indicate that most complications that arise during pregnancy, childbirth, or the puerperium occur during gestation, with higher rates among women aged 40 and over, preceded by the age group of 14 to 19 years, and these are highly preventable or treatable. Others may be present before pregnancy but worsen during pregnancy, especially if not treated as part of women’s health care or maternal health care (1,9,28,29).

Most of them are foreseeable, undoubtedly linked to insufficient education and cultural barriers of the population to the use of medical services, aggravated by inaccessibility to obstetric care and the limited quality of such care, in addition to the lack of knowledge and understanding of the problems that occur during pregnancy(30,31). The Latin America and the Caribbean region have health systems with limited capacity, with lower health financing, fewer hospital beds, lower satisfaction levels, and a lower doctor/patient ratio (32).

### Maternal Mortality and Covid-19

During the pandemic, the World Health Organization warned that pregnant women were at greater risk of developing more aggressive forms of COVID-19 and, in some cases, could progress to death (18). Physiological and immunological changes during pregnancy can lead to more complicated clinical events, causing fetal distress, premature rupture of membranes, premature births, and stillbirths (33). Takemoto comment that, for developing countries, high birth rates and limited health resources would result in an increase in maternal deaths (15).

Regarding care, there is suspicion of a higher risk of postoperative morbidity and mortality for women with COVID-19 undergoing cesarean section surgery (1,32,34). Regarding the cesarean section rate in the Dominican Republic, even without COVID-19, it is close to 58%, which is among the highest in the world, far from the 15% accepted by the WHO (35).

Unplanned pregnancies, deficiencies in prenatal care, the concentration of resources to address the coronavirus, and the large number of cesarean births are situations that affect the Dominican Republic and, along with other reasons, could worsen the Dominican reality regarding maternal mortality during the coronavirus pandemic(36). This leads to a hypothesis that COVID-19 intensifies the inequalities associated with maternal mortality.

### Social Determinants of Health and Covid-19

Health inequality are not limited to access to medical services or certain types of care. The social determinants model illustrates how social dynamics interplay to create social gradients in health (37). Housing, education, employment, geography, environment, and economic conditions directly or indirectly affect their well-being and health (18,37,38).

Contextual factors that create social hierarchies or stratifications in societies produce inequalities in maternal health (39–41). There is evidence that in the Dominican Republic, maternal mortality is higher in territories with higher levels of multidimensional poverty or in localities with lower levels of human development. Furthermore, the probability of a woman dying from causes related to pregnancy, childbirth, or the postpartum period in the most socially disadvantaged territories was about twice as high compared to the most socially advantaged territories (8).

Within the structural determinants, factors such as economic level, race and ethnicity, religion, gender, education, and occupation are also important (41–43). Regarding intermediate health determinants, Hamal (41) point to factors in the community context that include: area of residence, family context (family structure, decision-making power, access to resources, marital communication), peer cohesion (support networks), factors of the individual context (biological, such as age, parity, weight, height, health status (infections and parasitic diseases), behavior (family planning, pre-/intra-/postnatal care, emergency obstetric care, harmful traditional practices), and psychosocial factors.

Also, the part corresponding to the health system encompasses factors such as: availability of health systems (family planning, pre-/intra-/postnatal care, emergency obstetric care, referral, blood), accessibility (distance, time, transportation, transportation cost, formal and informal payments for health services, medicines and supplements, opportunity costs, companions, bribes, distribution and location of health facilities), quality of care (previous experience, satisfaction with services and costs, quantity of personnel/competence, diagnosis and action management), and lastly other unknown or unpredicted factors (41,42).

In the pandemic crisis, the worsening of inequalities is largely due to containment policies used to overcome the crisis (44,45). Using the words of Giannopoulou and Tsobanoglou (46), limiting access to healthcare for a significant portion of the population, closing medical centers for in-person visits, postponing appointments, and providing medical and diagnostic assistance to non-priority individuals due to justified fear of infection in hospitals translates into a significant impact on some pathologies.

In the case of COVID-19, they indicate that social status largely determines the risk of the virus, accessibility to healthcare, and the effectiveness of confinement measures (47). Conversely, the virus tends to increase inequality by reducing the incomes or work capacity of those infected or those who have stopped working due to restrictions (48). The aforementioned corroborates that the situation has affected population groups at greater risk who face challenges in accessing healthcare services (49). In this particular case, women may be disproportionately affected by the crisis: their income levels are, on average, lower than those of men, their poverty levels are higher, and they are more likely to be exposed to domestic violence (32).

From a theoretical perspective, various proposals have been developed for the elaboration of conceptual models for studying the determinants of maternal mortality and severe maternal mortality and morbidity (5,38,50–52), including the use of SINAVE (53). The models address the problems by integrating potentially fatal complications associated with the reproductive process and its surveillance with variables representing socioeconomic contextual conditions at the municipal and departmental levels. The variety of data structures used in epidemiological studies and the availability of statistical procedures have increased the use of multilevel models. Some researchers like Maia (54) argue that multilevel models emerge as an alternative to traditional multivariate models by considering the intrinsic hierarchical nature of the data and analyzing the autocorrelation between risk factors at aggregation levels.

## MATERIALS AND METHODS

### Data

### Sistema Nacional de Vigilancia Epidemiológico (SINAVE)

We use data from the National Epidemiological Surveillance System (SINAVE), captured by the Dirección General de Epidemiología and coordinated by the Ministry of Public Health and Social Assistance (MSP). The population under surveillance in this database of severe maternal morbidity and maternal death (including late ones) consists of pregnant or postpartum women aged 10 to 49 years. In addition, for the notification of severe maternal morbidity, the following criteria are established: 1) diagnosis of eclampsia, hypovolemic shock, and septic shock. 2) need for transfusion (of 3 or more units of any blood product), ICU admission, and/or emergency surgical procedure. 3) organ dysfunction or failure: cardiac, vascular, renal, hepatic, metabolic, cerebral, respiratory, and/or coagulation (55). The database contains 80 variables related to the woman, childbirth, and healthcare institution. For specifications on the database, see (29).

According to ECLAC (56), since the implementation of SINAVE, there has been a significant improvement in coverage of maternal deaths. Over the years, maternal mortality has been limited by problems of underreporting, either due to omission, errors in recording causes of death (56), or differences in province-to-province coverage (57).

We conducted an analysis of data completeness and validation. Additionally, we corrected the SINAVE database based on the REDENAF coverage. Between the years 2015 and 2021, a total of 14,655 cases of severe maternal morbidity and maternal mortality were reported.

### Maternal Death Classification and COVID-19 Infection Classification

The causes of death during pregnancy, childbirth, and the postpartum period, or late maternal deaths, according to ICD-10, are listed in Table 1 (4,23). These are subdivided into three categories: direct obstetric deaths, indirect obstetric deaths, and late obstetric deaths.

**Table 1.**
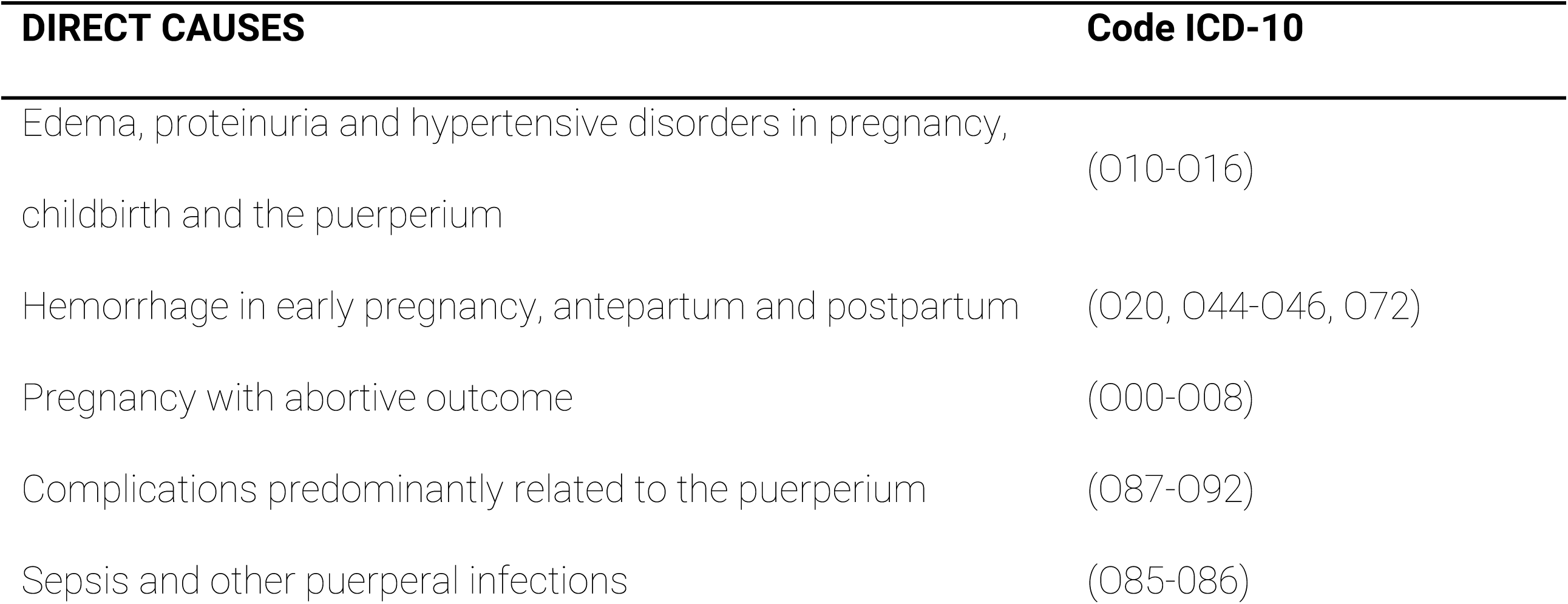
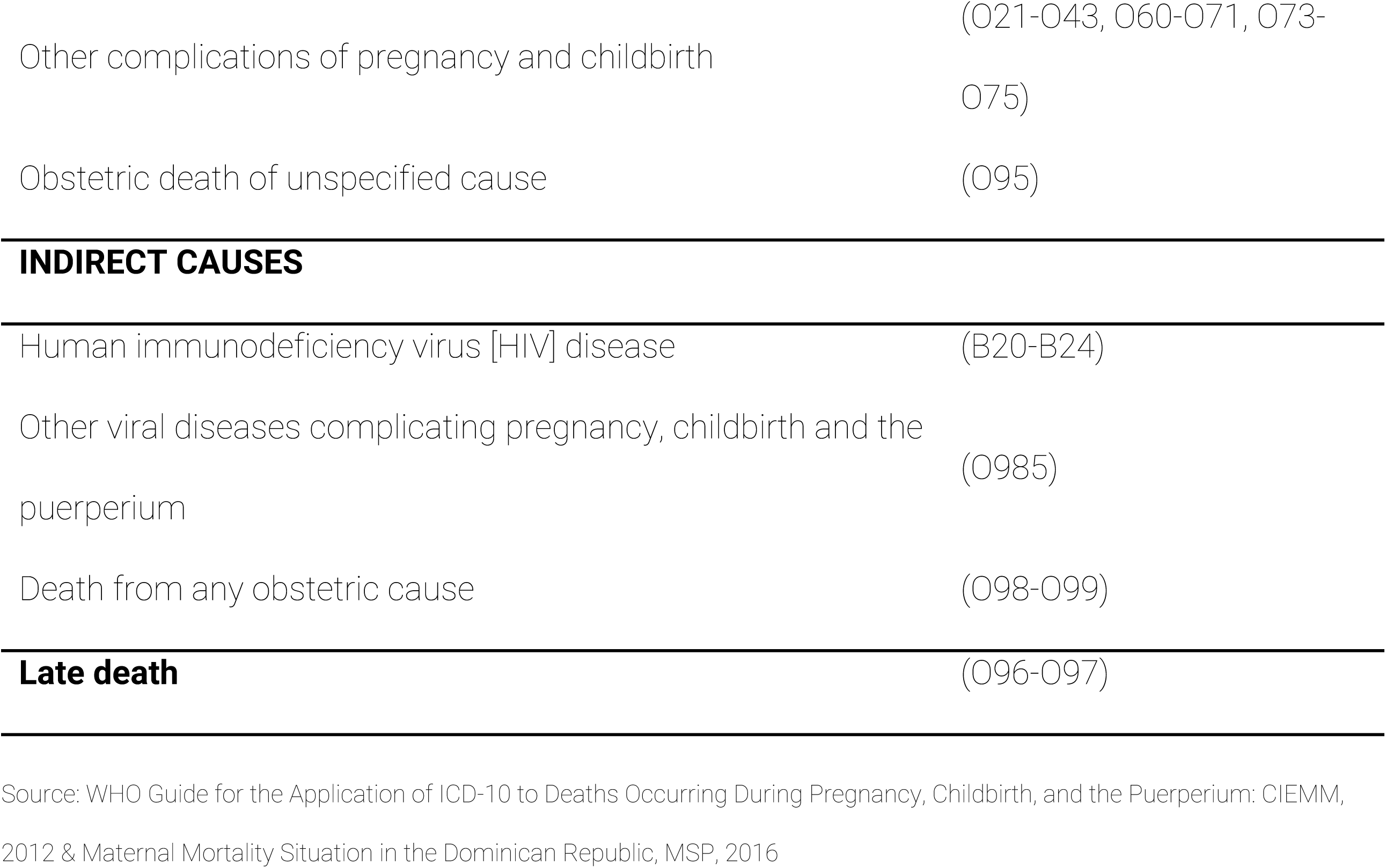
Groups of basic causes of death during pregnancy, childbirth, and the puerperium by ICD-10 codes.

In the Dominican Republic, there have been more than 659,761 cases of morbidity and over 4,384 deaths from COVID-19 infectious respiratory disease as of January 21, 2023 (Johns Hopkins Coronavirus Resource Center, 2023), with a mortality rate close to 39.7 per 100,000 inhabitants (58). The initial declaration of a health emergency by the WHO and the subsequent review of the pandemic required the creation of a new specific code for Covid-19, U07.1 (59). If during pregnancy, childbirth, or the puerperium, a woman is admitted (or has a medical care episode) for a condition due to COVID-19, it should be registered as the main diagnosis, with a code from the subcategory O98.5 "Other viral diseases complicating pregnancy, childbirth, and the puerperium", followed by the code U07.1. Codes related to COVID-19 always take priority in sequence (59).

### Live Births

Information regarding live births is available on the website of the Ministry of Public Health and Social Assistance of the Dominican Republic. These data comprise an annual consolidated report from three registration sources: the Weekly Report of Syndromes, Diseases, and Events of Mandatory Notification (EPI-1), the specialized services production form (67th), and the online birth registry database, resulting in a single consolidation for each establishment depending on the completeness of each source (55).

## METHODS

In order to estimate the Severe Maternal Morbidity Ratios (SMMR) and the Maternal Mortality Ratios (MMR) for the period 2015-2021 by cause of death (direct, indirect, and specific causes of the ICD), descriptive analyses were performed. Cases of Severe Maternal Morbidity (SMM) are identified as women who entered the surveillance system during pregnancy, childbirth, or the puerperium and left the system under the condition of "alive"; for Maternal Mortality (MM) cases, women in the "deceased" condition are considered. The main objective was to verify if there was a change in the observed behavior starting from 2020.

### Maternal Mortality Ratio

The Maternal Mortality Ratio (MMR) is the most commonly used measure of maternal mortality and is described as the relationship between the number of deaths due to maternal causes and the number of live births in the same period of time. To calculate it, late maternal deaths are excluded from the numerator. For the estimation of the MMR, maternal deaths of women aged 10 to 49 years were used for each year from 2015 to 2021 as the numerator, and live births from the same period were used as the denominator. Expressed per 100,000 live births, it measures the risk of death due to obstetric reasons once a woman becomes pregnant.

It is worth noting that for the calculation of this indicator, maternal deaths captured by SINAVE were used without correction, due to the good coverage and quality of SINAVE information. Literature indicates that there is no need to use a correction factor in provinces where the death is well investigated, such as in Belo Horizonte, for example (60). We also calculated the adjusted MMR (MMR_adj_) using a correction factor of 2.46% for maternal deaths in the numerator (representing the average percentage of non-coverage of SINAVE relative to RENADEF). The last is considered a more conservative ratio.

### Severe Maternal Morbidity Ratio

To estimate Severe Maternal Morbidity (SMM) in the Dominican Republic between 2015 and 2021, the SMM cases in women aged 10 to 49 years for each year observed in the period was divided by the number of live births in the period, according to the WHO guidelines (61). For this indicator, similar to the Maternal Mortality Ratio (MMR) indicator, no adjustment was made to exclude births of women older than 49 years from the denominator due to their low proportion.

### Proportion of maternal deaths due to a specific cause

The proportion of deaths due to a specific cause refers to the incidence of a death due to a specific cause as percentage of all maternal deaths. For the calculation of the proportion of deaths due to a cause, the number of direct and indirect obstetric deaths were used as the numerator, divided by the total number of maternal deaths (62).

### Maternal Mortality Ratio by specific cause

The Maternal Mortality Ratio by specific cause corresponds to the number of maternal deaths due to a specific cause per 100,000 live births.

### Mixed effect models

To examine the sociodemographic inequalities to which women in pregnancy, childbirth, or the postpartum period were exposed before and during the Covid-19 pandemic, we used individual level data from SINAVE, which contains cases of extreme maternal morbidity and maternal mortality. Of the total of 14,655 complete case records, 1,431 (9.8%) presented the condition of interest (death), considered as the dependent variable. Womeńs characteristics were used as explanatory variables while contextual variables were aggregated at the level of each of the provinces of residence of women, constructed using the databases of the National Multiple-Purpose Household Surveys (63) conducted by the National Statistics Office of the Dominican Republic. Contextual variables are proportions calculated with women aged 10 to 49 years as the denominator in each province. A summary can be seen in Table 1.

Multilevel models were adjusted with the purpose of controlling the effect of observations in each of the 32 provinces (intracluster correlation). For this, mixed effects models with a logit link function (due to the dichotomous nature of the dependent variable) were performed, placing contextual variables and the year receiving health care as random effects, while individual-level variables were placed as fixed effects in the model. Additionally, interactions between the year of receiving care and residence in a border province were conducted under a multiplicative effect interaction. To obtain risk estimates (OR), the estimated coefficients from the model were exponentiated, and a level of statistical significance of 0.05 was assumed. R Studio software version 4.2.1 was used to carry out the analyses described.

## RESULTS

Severe Maternal Morbidity Ratio (SMMR)

In the period between 2015 to 2021, 13,224 cases of extreme maternal morbidity were registered in the National Epidemiological Surveillance System (SINAVE), involving women aged 10 to 49 years residing in the Dominican Republic.

Figure 1 shows the numbers of SMM cases and the SMMR by year. It can be observed that between 2015 and 2017, there appears to be a slightly declining pattern, with values of 9.5, 7.8, and 7.6 cases per 1,000 live births, respectively. Starting from 2018, it increased to 10.4 per 1,000 live births and followed an increasing trend in 2019 (12.1 per 1,000 live births). In 2020, the indicator seems to decrease, showing a ratio of 10.4 per 1,000 live births; however, what we could be witnessing is a decline in admissions of pregnant women and postpartum women, caused by the COVID-19 pandemic. As for 2021, a significant difference in the maternal morbidity ratio can be observed, which recorded a ratio of 16.5 per 1,000 live births.

**Figure 1:**
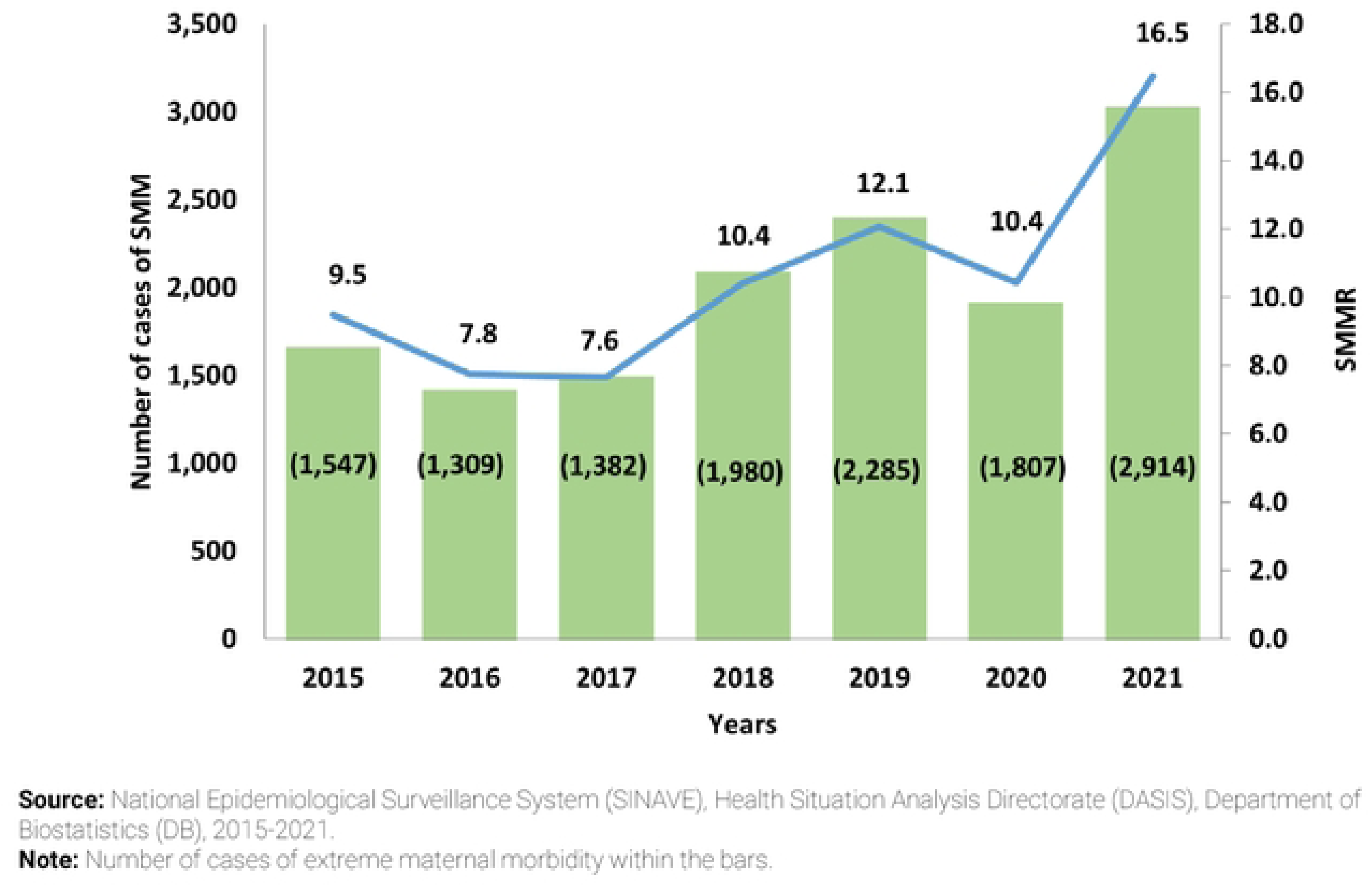
Severe maternal morbidity ratio and number of cases of severe maternal morbidity, Dominican Republic, 2015-2021.

### Maternal Mortality Ratio (MMR)

During the period 2015 to 2021, a total of 1,325 maternal deaths were recorded in the National Epidemiological Surveillance System (SINAVE), corresponding to women aged 10 to 49 residing in the Dominican Republic.

Figure 2 shows the changes in maternal mortality during the period 2015-2021. The initial years did not present significant changes, with the MMR in 2015 being 115.3 maternal deaths per 100,000 live births, followed by an MMR of 102.5 per 100,000 live births. For the following two years, higher rates were recorded, specifically in 2017 with an MMR of 111.3 maternal deaths per 100,000 live births and 2018 with an MMR of approximately 110.0 maternal deaths per 100,000 live births. Regarding the year 2019, it began to show a decline with an MMR of 91.4 per 100,000 live births, representing a decrease in the ratio of 18.6% compared to 2018.

**Figure 2.**
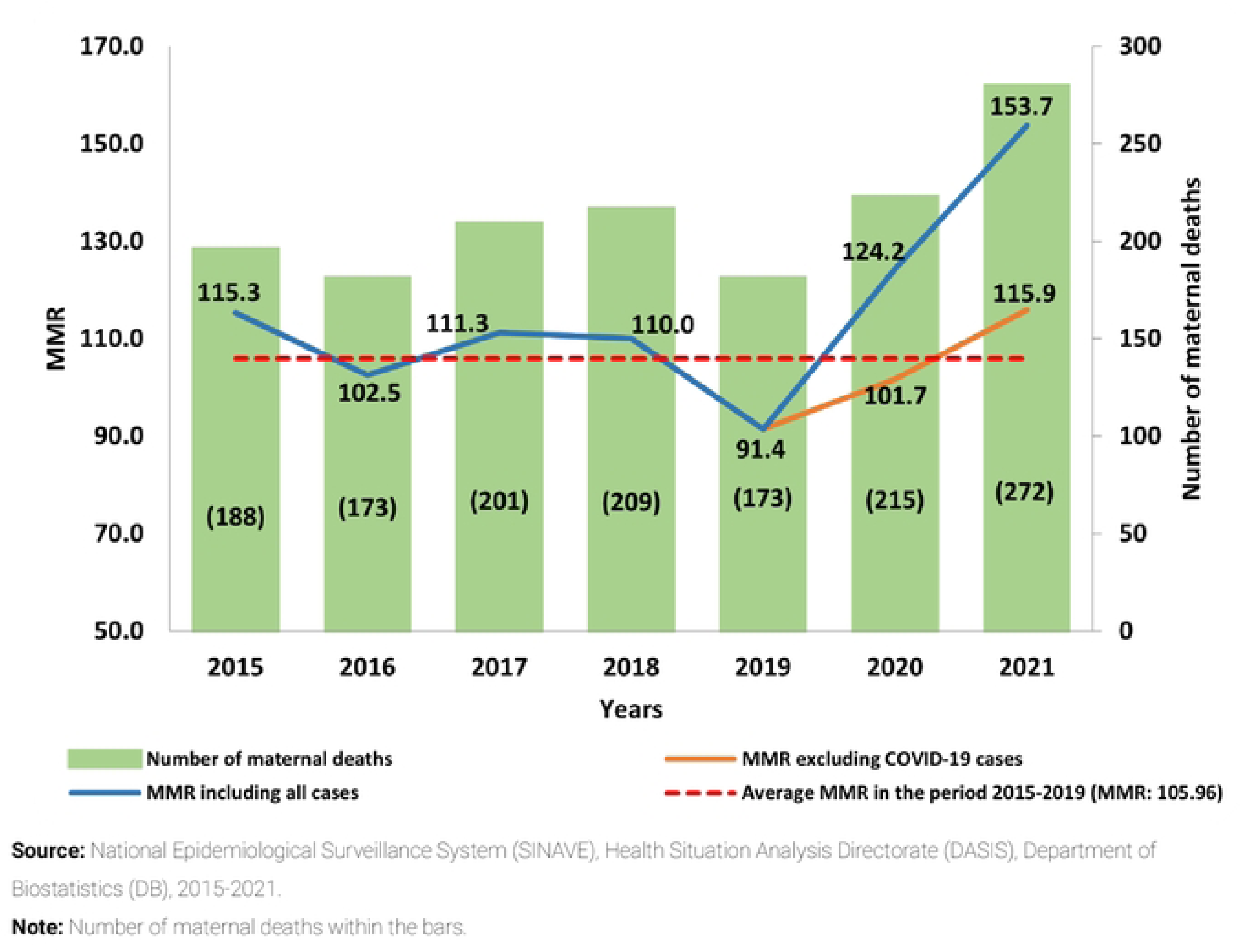
Maternal Mortality Ratio, Dominican Republic, 2015-2021.

Additionally, we can highlight in this graph the trend line with two branches that diverge in the years 2020 and 2021 showing a simulation. In blue, the maternal mortality ratio for the period with maternal deaths including COVID-19 cases in 2020 and 2021 is presented, while in orange, starting from 2019, are the MMRs for 2020 and 2021, but in this case, excluding maternal deaths due to COVID-19.

Regarding the MMR excluding COVID-19 cases (orange line, Figure 2), based on a counterfactual scenario, in the years 2020 and 2021, there would have been an MMR of 101.7 and 115.9 maternal deaths per 100,000 live births, respectively, values that would have been close to the average MMR in the period 2015-2019 (dotted red line). In other words, the evolution of the MMR for the period 2015-2021, without COVID-19 cases, pointed to a stagnant pattern, as oscillations around similar values were observed.

Exiting the counterfactual scenario, including all reported cases (blue line, Figure 2), a notable increase in the MMR for the years 2020 and 2021 compared to previous years is observed, with this being 124.2 and 153.7 maternal deaths per 100,000 live births, respectively, indicating a significant rise in the indicator for the latter year.

### Causes of Maternal Deaths

The analysis of the distribution of maternal mortality according to causes of death in the for the period 2015-2021 is presented in Figure 3. The following characteristics are observed: direct obstetric causes are responsible for more than 70% of maternal deaths from 2015 to 2020, except for the year 2021, which registred 62.5% of maternal deaths. On the other hand, indirect obstetric causes remained constant from 2015 to 2017, showing values close to 27%, while in the years 2018 and 2019 they decreased to 23.4% and 19.7% respectively. Regarding 2020 and 2021, there was a considerably high increase of 29.3% and 37.5% respectively.

**Figure 3.**
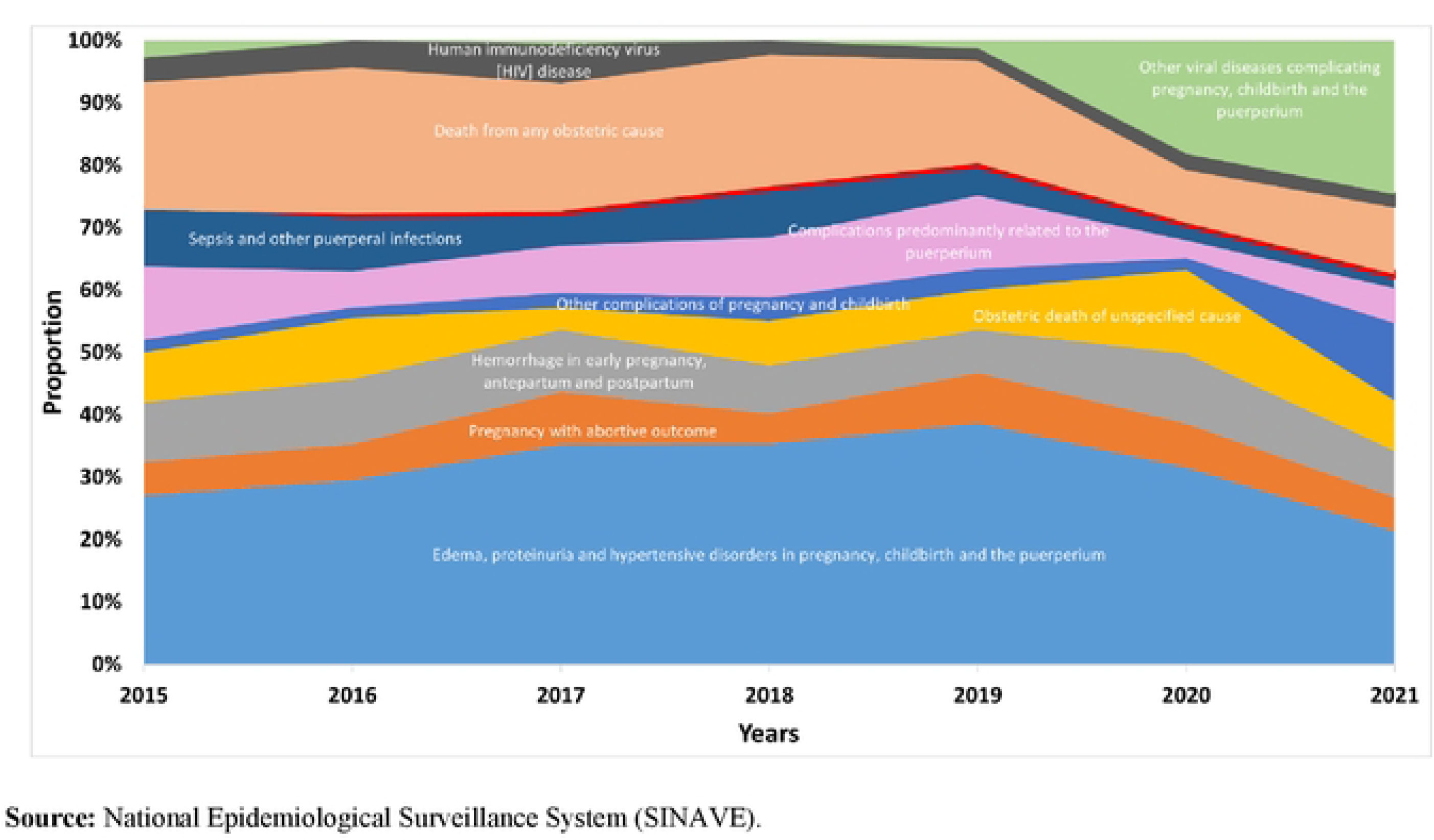
Percentage distribution of maternal mortality according to causes of Maternal deaths per year, Dominican Republic, 2015-2021.

Considering the literature regarding obstetric transition, there is a predominance of direct obstetric causes in all years, the most easily preventable. Within direct maternal causes, the group of edema, proteinuria and hypertensive disorders in pregnancy, childbirth and the puerperium predominates, ranging from 21.3% to 38.7%, followed by antepartum, intrapartum, and postpartum hemorrhage, and other complications of the postpartum period.

The role of pre-existing conditions is highlighted in indirect mortality, which is more difficult to reduce. In this regard, this subcategory presents proportions of 2.7% in 2015, 0.5% in 2017, and 1.2% in 2019; no cases of maternal death under this cause of death were recorded in 2016 and 2019. However, for the years corresponding to the COVID-19 pandemic, the proportion of cases under this category corresponds to 18.1% and 24.6% for the years 2020 and 2021, respectively. In numbers, in 2020, 39 cases of maternal deaths were registered under this cause, while in 2021, 67 cases of maternal deaths classified as COVID-19 were recorded.

The MMR due to direct obstetric causes was of 84 maternal deaths per 100,000 live births in 2015 (Table 2). In comparison, in 2021, the number was 96.1 maternal deaths per 100,000 live births. During the period, there was an increase of 12.1 maternal deaths due to direct obstetric causes per 100,000 live births. Maternal mortality is high regardless of COVID-19. The MMR due to "other viral diseases complicating pregnancy, childbirth, and the puerperium," which in the years before the COVID-19 pandemic (2015–2019) had shown a maximum value of 3.1 deaths per 100,000 live births (in 2015). In 2020, it reached 22.5 deaths per 100,00 live births, peaking in 37.9 in 2021.

**Table 2.**
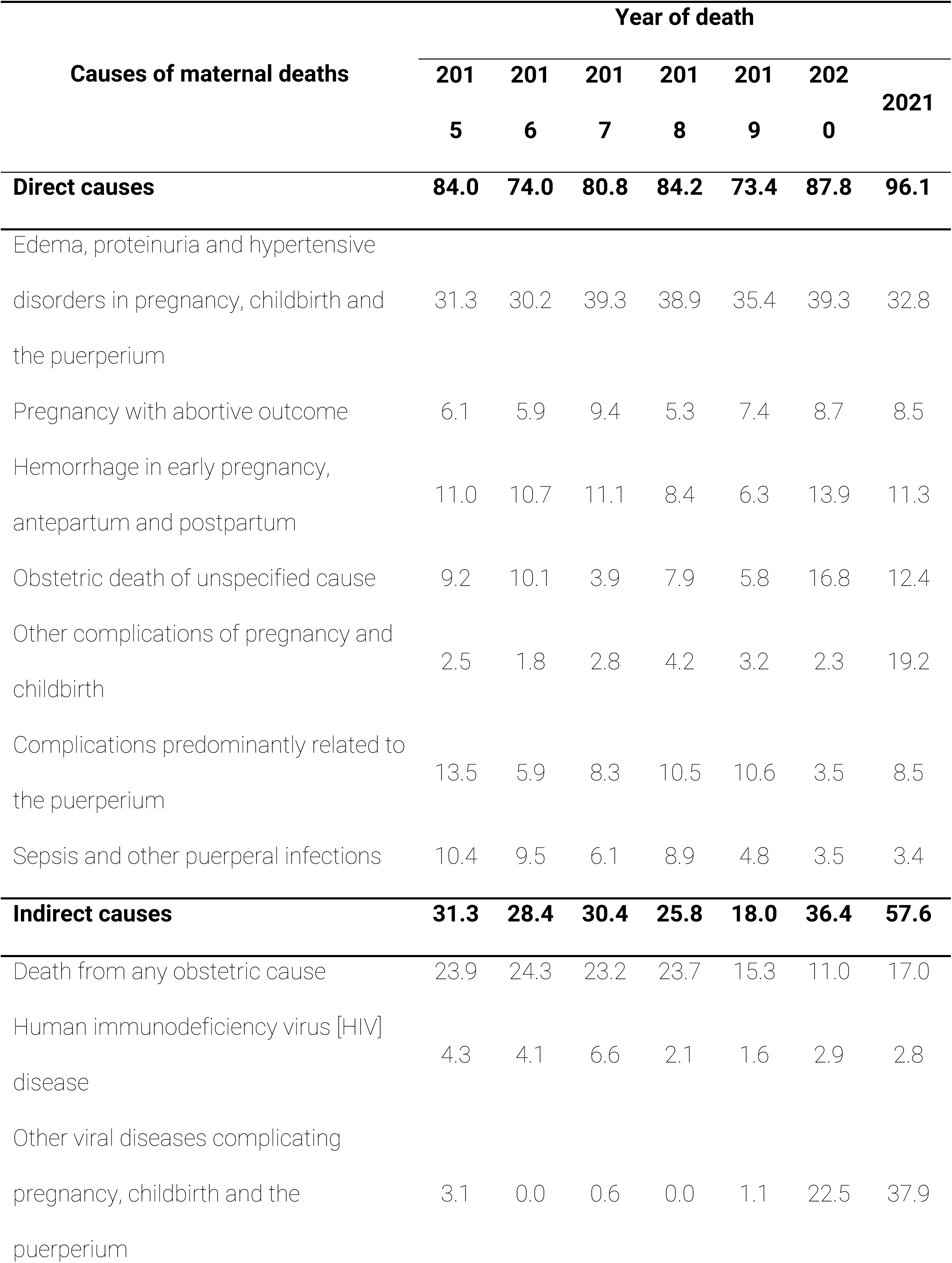
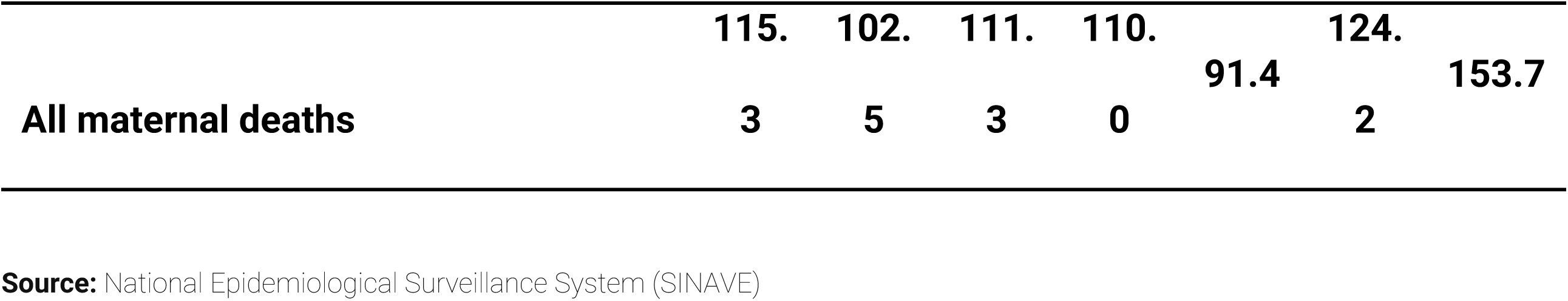
Maternal Mortality Ratio by Specific Cause per Year, Dominican Republic, 2015-2021.

Additionally, the MMR for "antepartum, intrapartum, and postpartum hemorrhage" and "obstetric death of unspecified cause" show an increase in 2020 of 13.9 and 16.8, respectively. The MMR for other complications of pregnancy and childbirth, which in the period from 2015 to 2020 showed values between 1.8 and 4.2, however, increased to 19.2 deaths per 100,000 live births in 2021.

### Contextual sociodemographic of maternal deaths

Figure 4 presents the spatio-temporal evolution of the maternal mortality ratio for each of the provinces in the Dominican Republic for the period 2015 to 2021, in addition to the period average. Thus, despite the Dominican Republic presenting a considerably high maternal mortality ratio, the national average tends to conceal the heterogeneity of the health situation within the country (8).

**Figure 4.**
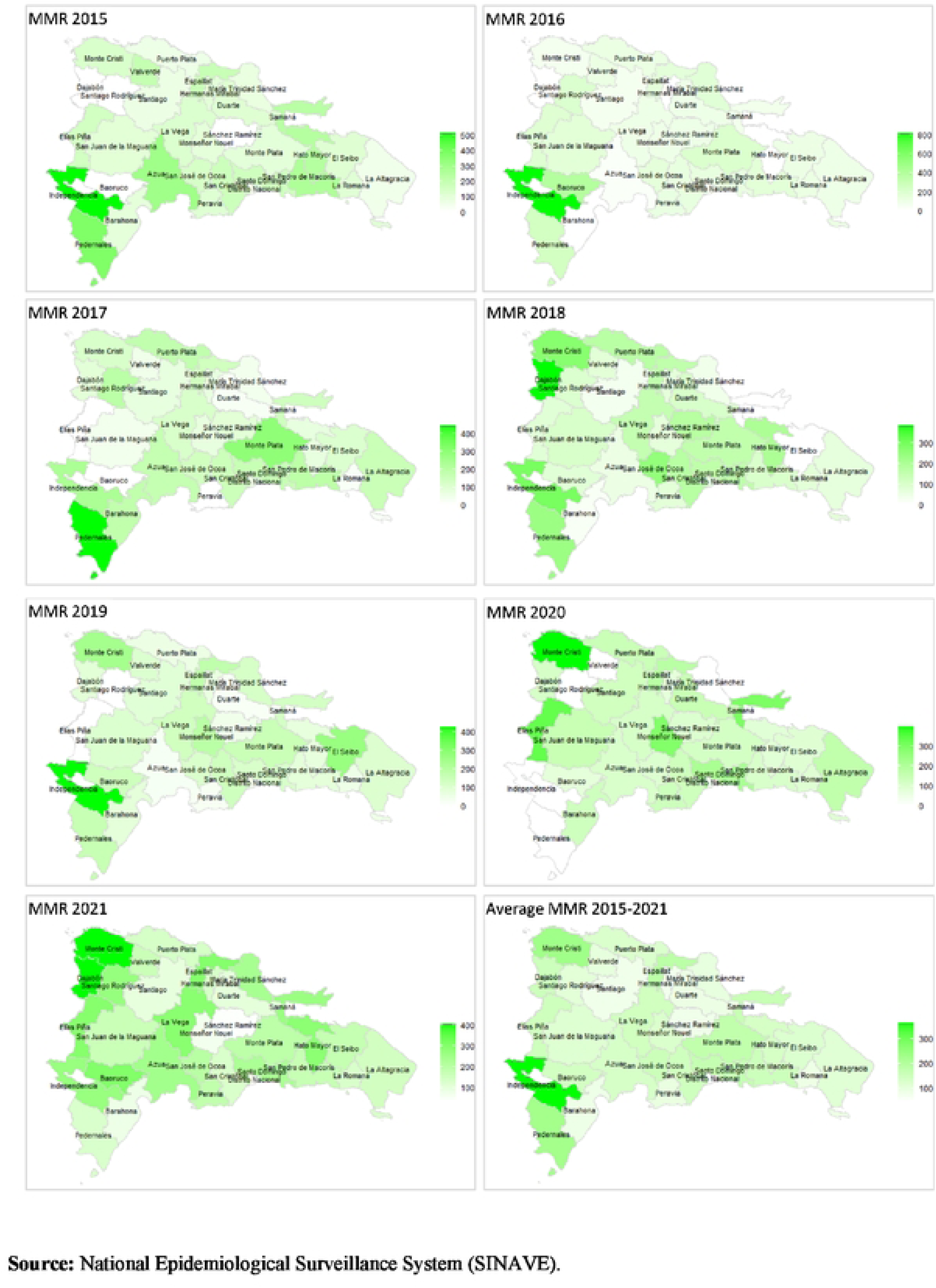
Maternal Mortality Ratio by province of residence, Dominican Republic, 2015-2021.

The analysis by provinces allows identifying the locations with the highest ratios and, based on this, prioritizing them for further analysis and identifying possible associated factors. The national average of the maternal mortality ratio for the period 2015-2021 shows that the provinces Independencia (MMR: 364/100,000 live births), Monte Cristi (MMR: 232.7/100,000 live births), and Pedernales (MMR: 227/100,000 live births) showed the highest levels of this indicator. On the other hand, the provinces Distrito Nacional (MMR: 44.1/100,000 live births), Hermanas Mirabal (MMR: 74.6/100,000 live births), and Santiago (MMR: 77.8/100,000 live births) presented the lowest levels in relation to the maternal mortality ratio.

In Table 3, some sociodemographic characteristics of women who presented conditions of severe maternal mortality or morbidity during the period 2015-2021, captured by SINAVE, are shown. In general terms, around 10% of these women died due to maternal mortality during the period.

**Table 3.** Sociodemographic Characteristics of Women aged 10 to 49 who Presented Conditions of MM or SMM, Dominican Republic, 2015-2021.

| Characteristics | Maternal deaths |  | Total<br>N=14,655 (100%) |
| --- | --- | --- | --- |
|  | No | Yes |  |
|  | N=13,224 (90.2%) | N=1,431 (9.8%) |  |
| Age group, n (%) |  |  |  |
| 10_19 | 3001 (22.7%) | 202 (14.1%) | 3203 (21.9%) |
| 20_29 | 6409 (48.5%) | 641 (44.8%) | 7050 (48.1%) |
| 30_39 | 3434 (26.0%) | 513 (35.8%) | 3947 (26.9%) |
| 40_49 | 380 (2.9%) | 75 (5.2%) | 455 (3.1%) |
| **Pregnant, n (%)** |  |  |  |
| No | 4172 (31.5%) | 398 (27.8%) | 4570 (31.2%) |
| Yes | 9051 (68.4%) | 1032 (72.1%) | 10083 (68.8%) |
| Missing | 1 (0.0%) | 1 (0.1%) | 2 (0.0%) |
| **Residing in border provinces, n (%)** |  |  |  |
| No | 12530 (94.8%) | 1352 (94.5%) | 13882 (94.7%) |
| Yes | 692 (5.2%) | 78 (5.5%) | 770 (5.3%) |
| Missing | 2 (0.0%) | 1 (0.1%) | 3 (0.0%) |
| **Nationality, n (%)** |  |  |  |
| Dominican | 10837 (81.9%) | 1103 (77.1%) | 11940 (81.5%) |
| Haitian | 2352 (17.8%) | 324 (22.6%) | 2676 (18.3%) |
| Others | 35 (0.3%) | 4 (0.3%) | 39 (0.3%) |
| **Time of care within less than 24 hours, n (%)** |  |  |  |
| No | 4199 (31.8%) | 636 (44.4%) | 4835 (33.0%) |
| Yes | 9025 (68.2%) | 795 (55.6%) | 9820 (67.0%) |
| **Mobility for seeking care, n (%)** |  |  |  |
| No | 9102 (68.8%) | 936 (65.4%) | 10038 (68.5%) |
| Yes | 4120 (31.2%) | 494 (34.5%) | 4614 (31.5%) |
| Missing | 2 (0.0%) | 1 (0.1%) | 3 (0.0%) |
| **Infected with Covid-19 during the pandemic\*** |  |  |  |
| No | 4,472 (94.7) | 388 (79.2) | 4,860 (93.3) |
| Yes | 249 (5.3) | 102 (20.8) | 351 (6.7) |
| **Comorbidities** |  |  |  |
| No | 10398 (78.6%) | 1006 (70.3%) | 11404 (77.8%) |
| Yes | 2826 (21.4%) | 425 (29.7%) | 3251 (22.2%) |
| **Year of attention** |  |  |  |
| 2015 | 1547 (11.7%) | 190 (13.3%) | 1737 (11.9%) |
| 2016 | 1309 (9.9%) | 172 (12.0%) | 1481 (10.1%) |
| 2017 | 1382 (10.5%) | 203 (14.2%) | 1585 (10.8%) |
| 2018 | 1980 (15.0%) | 208 (14.5%) | 2188 (14.9%) |
| 2019 | 2285 (17.3%) | 168 (11.7%) | 2453 (16.7%) |
| 2020 | 1807 (13.7%) | 220 (15.4%) | 2027 (13.8%) |
| 2021 | 2914 (22.0%) | 270 (18.9%) | 3184 (21.7%) |

**Source:** National Epidemiological Surveillance System (SINAVE)
**Note:** \*for the denominator, cases for 2020 and 2021 were used.

Around 2 out of 10 women were of Haitian nationality (18.3%), with this proportion being even higher among women who lost their lives due to maternal causes (22.6%). It is worth noting that, while other nationalities are observed, this proportion is relatively low, reaching less than 1% in the data. Among all female inhabitants of the Dominican Republic, it is estimated that only 6.2% are Haitian migrants, and 0.7% are of other nationalities (64). Among live births, 13% are children of Haitian women, and 0.9% are children of women of other nationalities (65).

In Table 3 also, clinical and health care-related characteristics are showed. Regarding comorbidities present in this population, it was reported that 22.2% of these women had some comorbidity, with this proportion considerably higher (29.7%) in the group of women who died.

In general terms, the average length of stay was 1.30 days (SD = 4.54); however, the average length of stay for women who died was 3.19 days (SD = 8.35). It is observed that nearly 7 out of 10 women (67.0%) received care within the same day; however, this proportion decreases in the group of women who died (close to 5 out of 10 women, or 55.6%).

Regarding mobility in seeking care, peregrination, we can observe that 3 out of 10 women (31.5%) received care in a province different from the one reported as their province of residence, a proportion that in the group of women who died, turned out to be slightly higher (34.5%). During the years 2020 and 2021, in general terms, 6.7% of these women reported having been infected; however, this proportion is 3.1 times higher in the group of women who died (20.8%).

In the adjusted models, as can be seen in Table 4, comparing to the group age 10-19, the higher the womeńs age, the highest the chance of mortality. Other individual characteristics also increases the risk, such as being of Haitian origin (36.7%, p-value < 0.001) being pregnant (17.8%, p-value = 0.024) and presenting comorbidities (76,2%, p-value < 0.001). Regarding characteristics related to the health care system, receiving care on the same day symptoms began is considered a protective factor against maternal mortality (p-value = 0.024), reducing the risk of death by 33.2%.

**Table 4.**
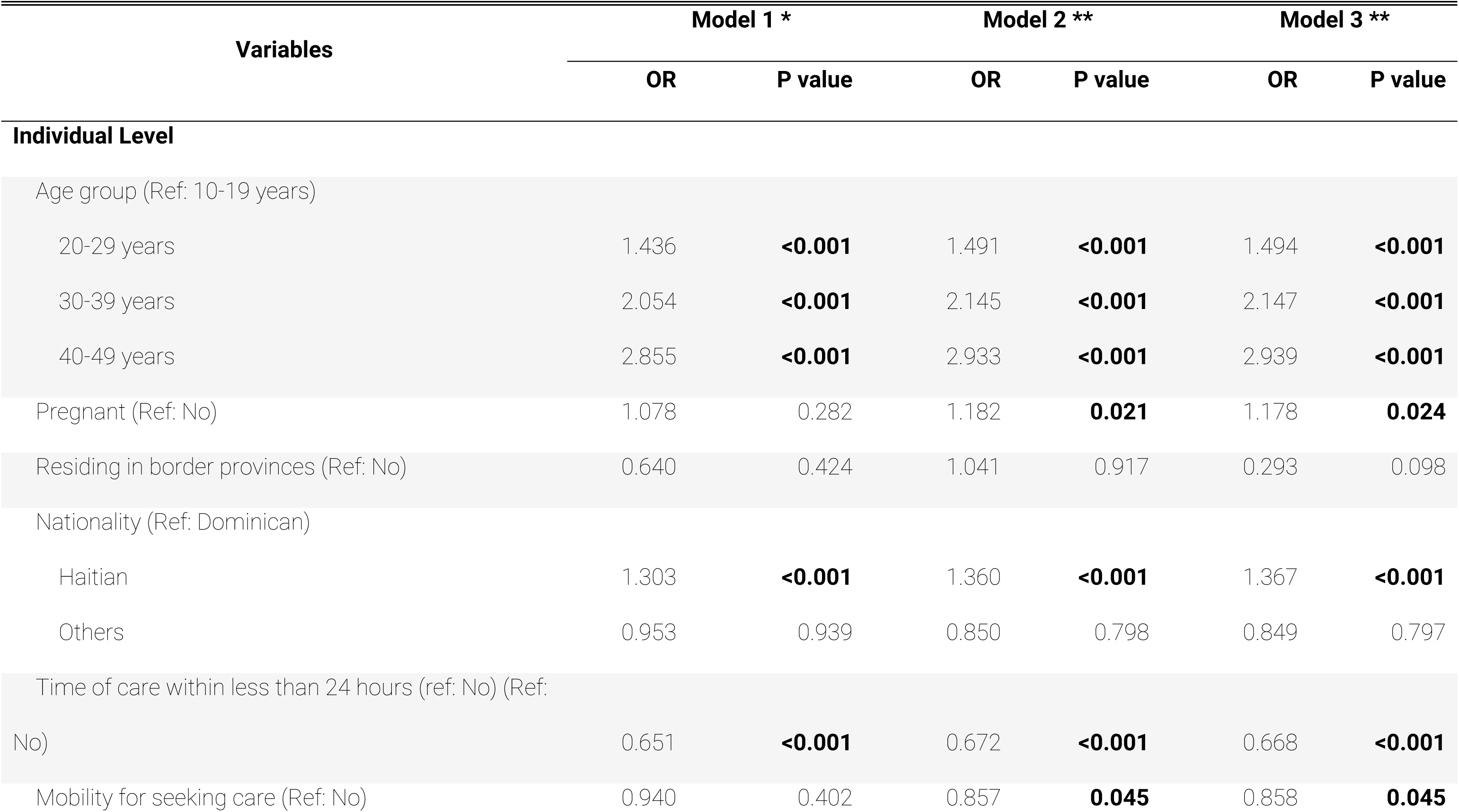
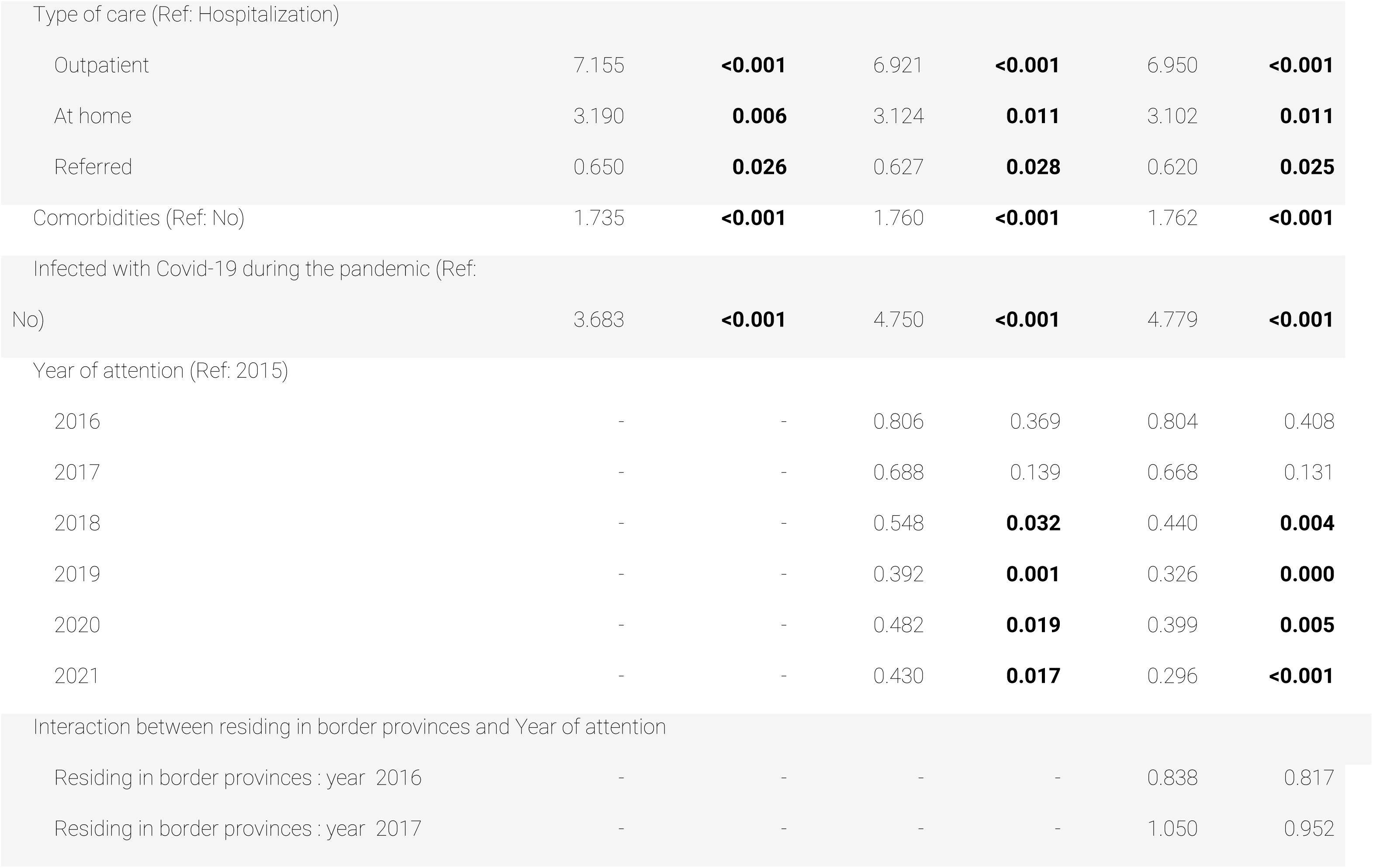
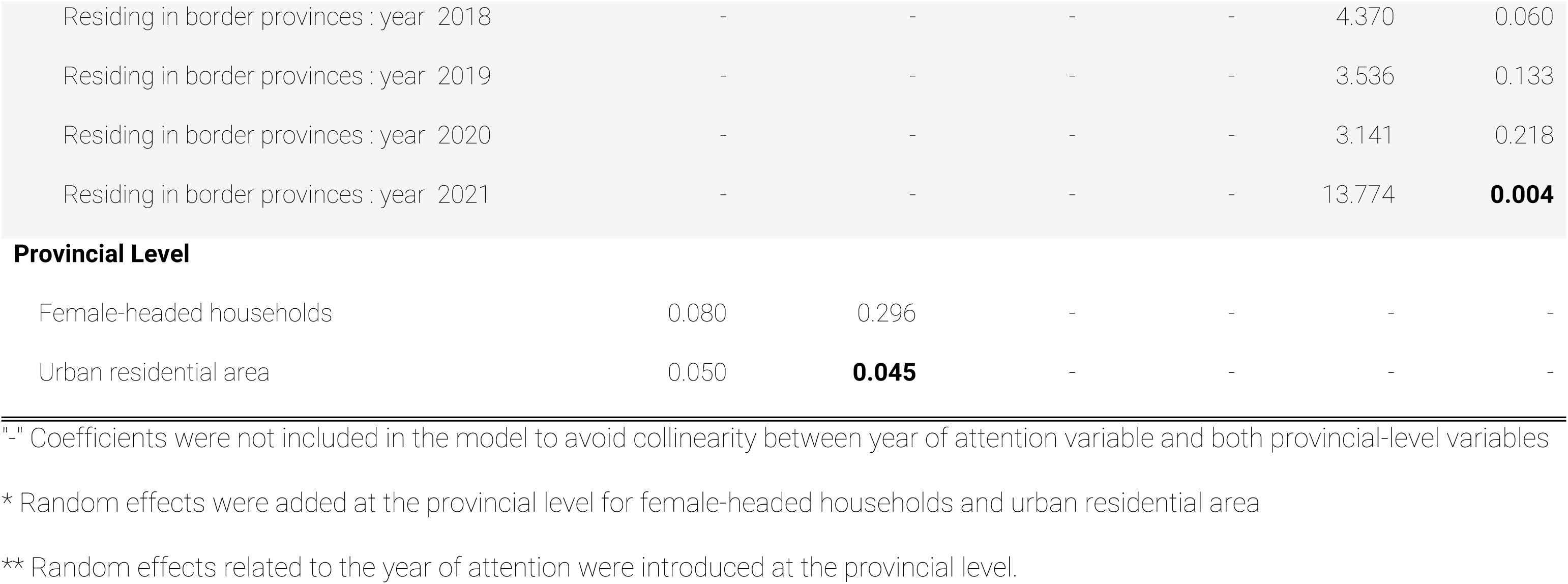
Multivariate model of association for maternal mortality using severe maternal morbidity cases as reference.

When comparing women who received outpatient care with women who got hospitalized, the former have a 6.9 times higher risk of dying (p-value < 0.001). Similarly, receiving care at home is also considered a risk factor for mortality (Odds: 3.10 p-value = 0.006) compared to women who were hospitalized. Conversely, receiving care through referrals (women who were already hospitalized or received care at another facility) is considered a protective factor against mortality (p-value = 0.026), reducing the risk of maternal death by 37.8%. Mobility for seeking care is associated with less maternal mortality (p-value = 0.045), reducing the risk of maternal death by 14.2%.

With regard to characteristics associated with the context of the COVID-19 pandemic, women in SMM condition who reported being infected with the virus have a 4.8 times higher risk of dying from maternal death than women who were not infected (p-value < 0.001). Taking 2015 as a reference year, all years (2016 to 2021) demonstrate fewer chances of observing maternal mortality, being significant for the years 2018 (p-value = 0.032), 2019 (p-value < 0.001), 2020 (p-value < 0.049), and 2021 (p-value < 0.072). This can be translated as an improvement in care; however, this improvement in care was not general for all women, as observed in the interactions of model 3. In this adjusted models, the provinces located in the southwest region of the Dominican Republic, the ones that border the Republic of Haiti and are the ones with the worst socioeconomic conditions, are associated with a 13 times increase in the odds of dying in the year 2021 (p-value = 0.002) and indicating a 13.8 times higher risk for women residing in these border provinces compared to women who do not reside in these provinces.

## Discussion and Conclusion

The first objective of this study was to estimate severe maternal morbidity ratios and maternal mortality ratios for the period 2015-2021, verifying if, starting from 2020, there was a change in the observed behavior for the period. We find that during the years 2015 to 2019, severe morbidity showed a slightly pronounced pattern of increase, which could be attributed to the improvement in the surveillance protocol of the SINAVE; however, in 2020, the indicator seems to decrease, showing a ratio of 10.4 for every 1,000 live births. The first hypothesis suggests a decrease in the income of pregnant women and puerperal women, caused by the COVID-19 pandemic, which according to Rivera Alvarado (66), impacted access to healthcare services leading to a parallel crisis in sexual and reproductive health and a consequent exacerbation of social inequalities.

In the same vein, OPS (16) stated that COVID-19 caused devastating impacts on women, pregnancy care, and newborn care, with the virus interrupting services in almost half of the countries in the Americas. At the same time, it indicates that pregnant women are more vulnerable to respiratory infections such as COVID-19 and that if pregnant women become ill, they tend to develop more severe symptoms.

The second scenario regarding the decrease in SMMR in 2020 could be due to a change in the detection of severe maternal morbidity cases. Recent research has shown evidence that problems arose in the notification and properly conducted diagnosis during the first year of COVID-19, causing a disruption in the 2020 data (67). In 2021, the indicator recorded a ratio of 16.5 for every 1,000 live births.

Regarding the results of the estimates of the Maternal Mortality Ratio for the period 2015 to 2021 in the Dominican Republic, until the year 2018, the MMR did not present significant changes, that is, the evolution of this indicator seemed stagnant, except for the year 2019, which decreased to an MMR of 91.4 for every 100,000 live births. The MMR for 2020 and 2021 compared to previous years shows a significant increase of 124.2 and 153.7 maternal deaths per 100,000 live births, respectively, which can be directly linked to the COVID-19 pandemic. This increase in MMR associated with COVID-19 in the years 2020 and 2021 has also been recorded in other countries in Latin America and the Caribbean, such as Brazil. According to Valongueiro (68) in Rio de Janeiro, preliminary information for 2021 showed an MMR of 201.7 maternal deaths per 100,000 live births, for an increase in MMR between 2019 and 2021 of 150%.

One way to measure the impact of the pandemic is to analyze excess mortality, that is, the level of mortality at a given time compared to recent trends or, in this case, the expected value (67). Adopting the practices carried out by Lima (67), when comparing the number of maternal deaths for the years 2020 and 2021 with the average for previous years (2015–2019) (188.8 deaths), we have an excess of 13.9% and 44.1% of maternal deaths in 2020 and 2021, respectively. Repeating this exercise to estimate the excess deaths for general mortality (all people, not just obstetric), using the data of registered deaths (65), for the years 2020 and 2021, an excess in general mortality of 6.1% and 10.6% was recorded, respectively.

Direct obstetric causes account for more than 70% of maternal deaths from 2015 to 2020, except for the year 2021, which showed 62.5% of maternal deaths. On the other hand, indirect obstetric causes from 2015 to 2017 remained constant, showing values close to 27%, while in 2018 and 2019 they decreased to 23.4% and 19.7%, respectively. The proportion of deaths caused by the subcategory "Other viral diseases that complicate pregnancy, childbirth, and the postpartum period“ reached 18.1% and 24.6% for the years 2020 and 2021, respectively. Concerning 2020 and 2021, indirect causes, such as the viral infections, had a considerably high increase of 29.3% and 37.5%, respectively. Despite the increase in indirect obstetric causes, according to Say (61), severe hemorrhages (mostly postpartum) and complications during childbirth are among the main complications that cause 75% of maternal deaths. Throughout the period there was an increase of 12.1 maternal deaths due to direct obstetric causes per 100,000 live births, an increase that can be attributed to the disruptions that the healthcare system had due to the COVID-19 pandemic (67).

The results also show that womeńs age is considered a risk factor related to maternal mortality. The results also highlight the proportion of cases in adolescent women (21.9%), compatible with adolescent pregnancy rates in the Dominican Republic (63). At the other end of the age spectrum are women aged 40 to 49, representing about 3% of maternal and extreme maternal mortality cases. These results are related to those presented by Donoso and colleagues (69), which state that pregnancy at the extremes of reproductive age is a risk factor for maternal, perinatal, and infant morbidity and mortality.

Other findings are supported by the literature, such as the fact that complications or comorbidities may exist before pregnancy, but these worsen during gestation, particularly when not treated as part of women’s healthcare (9,18,20,28). The results also highlight the importance of providing immediate care (28) for improving the chances of survival.

Thinking about the role of the social determinants of health for the maintenance of health inequalities (39), especially when it comes to sexual and reproductive health, this paper adds to the existing evidence that maternal mortality is higher in territories with higher levels of multidimensional poverty or in localities with lower levels of human development. Patricio and Santos (8) show how the probability of a woman dying from causes related to pregnancy, childbirth, or the postpartum period in the most socially disadvantaged territories was around twice as high compared to the most socially favored territories. Our results revealed that being a resident of an international bordering area in 2021 multiplied the chances of dying by 13; and being of Haitian nationality carried out a 30.3% higher chance of incurring in maternal death compared to a woman of Dominican nationality. Knowing that economic level, race and ethnicity, religion, gender, education and occupation have an impact on health outcomes through intermediate determinants (41,70) leads to the assumption that there is an intersection of these factors in Haitian women, migrants, possibly of low income, residing in more disadvantaged localities. It is worth emphasizing that at the population level, the greatest migration received by the Dominican Republic is from Haitian citizens due to the proximity and vast social and economic differences that favor this situation (9). As the World Health Organization states, research on severe maternal morbidity can indicate the way to reduce high maternal mortality rates; however, more studies should be conducted that include social, economic, political, and health determinants in the Dominican Republic and more public policy is needed to address the sociodemographic and economic conditions of women in order to improve access to and use of health services.

The findings show that the pandemic brough up a catastrophic situation for maternal health. However, maternal mortality is high in the Dominican Republic regardless of COVID-19. In addition, there was an increase in poorly specified causes, so the numbers might be understimated. As the country has not been able to experience a significant reduction in maternal mortality without COVID-19 cases, it moves away from achieving the 2030 Agenda and the Sustainable Development Goals. (1)

## Data Availability

The data cannot be published because it is data from the registry of the Ministry of Public Health of the Dominican Republic, it has to be requested from the institution.

https://www.one.gob.do/publicaciones/2019/encuesta-nacional-de-hogares-de-propositos-multiples-enhogar-2018-informe-eneral/?altTemplate=publicacionOnline

